# A Global Health Quality Improvement Project: Enhancing Cervical Cancer Awareness and Screening in Nigeria

**DOI:** 10.64898/2026.06.09.26355311

**Authors:** Ilham Umar, Nabila Shehu, Nancy Nagib, Suraiyya Sulley, Zubeda Oyiza Idris-Saeed

## Abstract

**Background:** Cervical cancer remains a significant global public health challenge, ranking as the fourth most common cancer among women worldwide. According to The World Health Organization (WHO) 604,000 women were diagnosed with cervical cancer globally in 2020, with over 342,000 deaths amongst this group [1]. Despite its high mortality, cervical cancer is largely preventable through early detection and vaccination against human papillomavirus (HPV), which causes nearly all cases of cervical cancer [1,2]

In Nigeria, it is the second most common cancer among women in Nigeria and a leading cause of cancer-related deaths, with low screening rates exacerbating late diagnoses and poor outcomes [1]. Despite global commitments to elimination with Pap smear screening and HPV vaccination, less than 10% of women in Nigeria have undergone screening due to misconceptions, stigma, and limited awareness. Educational interventions may improve awareness and promote screening behaviors. This global health quality improvement (QI) project aimed to enhance cervical cancer awareness and increase Pap smear uptake at the Central Bank of Nigeria (CBN) Clinic in Abuja, Nigeria.

**Methods:** In November 2024, we conducted a health education intervention at the Central Bank of Nigeria (CBN) through a structured educational session for male and female CBN staff members. The session focused on cervical cancer prevention, risk factors, and screening guidelines. Additionally, cervical cancer awareness was raised via email, social media, and electronic bulletin board. Participants completed pre and post-interventions surveys assessing cervical cancer knowledge across 10 key items and demographic characteristics. Pap smear uptake was assessed using the CBN clinic records for three months before and after the intervention. Institutional approval was obtained from CBN and external institutional review board approval was not required.

**Results:** 188 participants attended the health education session with 124 survey responses (70 pre-event, 54 post-event). Participants were mostly women aged 30–39. Post-intervention, eight of ten survey questions showed improved knowledge, with five demonstrating statistically significant gains: understanding Pap smear frequency (*p*<.001), HPV infection prevention (*p*=.042), early symptoms of cervical cancer (*p*=.019), smoking as a risk factor (*p*=.002), and availability of Pap smears at the CBN clinic (*p*=.035). Pap smear uptake increased from 5 screenings in three months pre-intervention to 32 screenings in the three months post-intervention. Participants reported that the sessions provided a safe space to ask questions and address cultural myths and misconceptions.

**Conclusion:** This QI initiative demonstrates the positive impact of targeted health education in improving awareness and screening uptake. Recommendations include increasing awareness through public health talks, updating clinicians on current guidelines, and removing unnecessary barriers to HPV vaccination. These findings align with global health efforts to reduce cervical cancer mortality and underscore the potential of QI projects to improve health outcomes in resource-limited settings.

## Introduction

Cervical cancer (CxCa) remains a leading but preventable cause of cancer morbidity and mortality among women in low- and middle-income countries, driven largely by persistent infection with high-risk human papillomavirus (HPV) [1]. Over the past decade, the global policy environment has shifted from incremental control to an elimination agenda, anchored by the WHO targets for 2030: 90% of girls fully vaccinated with HPV vaccine by age 15, 70% of women screened with a high-performance test at ages 35 and 45, and 90% of women with cervical disease treated [1,2]. Yet coverage gaps in vaccination, screening, diagnosis, and timely treatment persist, especially across sub–Saharan Africa. Nigeria contributes a substantial share of the continental burden, with high incidence and mortality compounded by late presentation and low lifetime screening prevalence [4,3].

Against this backdrop, workplace clinics represent a strategic yet understudied entry point for accelerating secondary prevention. They convene a relatively stable population, offer predictable access to health communication channels such as email, bulletin boards and social media, and can integrate screening pathways into existing occupational health services. Evidence from low- and middle-income settings indicates that structured health education, invitation and recall systems, and streamlined screen and treat models can raise uptake and reduce loss to follow up [2,11]. However, implementation frequently stalls on practical barriers that include knowledge and stigma, affordability and time costs, limited trained providers, supply constraints for ablative therapy, and weak data systems for tracking invitations, results and recalls [7,9].

Quality improvement methods offer a pragmatic pathway to bridge these operational gaps. Using the Model for Improvement and Plan-Do-Study-Act (PDSA) cycles, multidisciplinary teams from different disciplines can test small changes such as improving demand creation, appointment flow, data collection, and same-day treatment capacity. From there, teams can refine those changes based on feedback from run charts [12,13]. In this manuscript, we report on a quality improvement initiative conducted at the Central Bank of Nigeria Clinic in Abuja that sought to increase awareness and Pap smear uptake through targeted education and multi- channel communication. The Introduction and Background that follow position the project within the elimination agenda, synthesise evidence on barriers and effective strategies in low- and middle-income countries and in Nigeria, and explain why a quality improvement approach in a workplace clinic is timely and policy relevant.

## Background and Literature Review

### Global elimination targets and technical standards

The WHO Global Strategy to Accelerate the Elimination of Cervical Cancer frames CxCa as a preventable disease amenable to primary prevention through HPV vaccination and to secondary prevention through screening and treatment of precancerous lesions [1]. The 2021 WHO guideline recommends HPV DNA testing as the preferred primary screening modality because of superior sensitivity and programmatic efficiency; visual inspection with acetic acid remains acceptable where HPV testing is not yet available [2]. Evidence supports screen, triage, and treat pathways with same day thermal ablation for eligible lesions to minimise loss to follow up; for women living with HIV, earlier initiation and shorter screening intervals are indicated[2,11].

### Nigeria’s epidemiology, service landscape, and recent policy shifts

Nigeria cervical cancer burden reflects historically low screening coverage and late-stage diagnosis. In 2023, estimates indicated 13,676 new cases and 7,093 deaths, with age standardised incidence and mortality among the highest in the region [4]. Despite national and professional society guidance endorsing HPV vaccination and secondary prevention, adult screening remains uneven, with public sector services often relying on visual inspection with acetic acid because of cost, supply chain and laboratory capacity limitations for HPV DNA testing [6,2]. In October 2023, Nigeria introduced HPV vaccination into routine immunisation with a first-round target of 7.7 million girls, one of the largest introductions in Africa, marking an upstream policy milestone that now needs to be matched by expanded adult screening and treatment capacity [5]. While overall cancer statistics continue to be updated, GLOBOCAN 2022 data similarly underline cervical cancer place among the top three cancers in Nigerian women [3].

### Determinants of low awareness and low screening uptake

Barriers at individual, interpersonal, community and health system levels consistently suppress screening demand and completion in Nigeria and comparable settings [8,7,9]. Knowledge gaps about HPV and cervical cancer, misconceptions about fertility or pain, and low risk perception limit intent to screen. Stigma and fatalistic beliefs deter health seeking. Interpersonal dynamics, including partner influence and the need for explicit spousal permission in some households, shape the ability of women to attend services. At community level, religious or cultural norms may restrict discussions of sexual health or deter pelvic examinations. On the supply side, costs, both direct and indirect, distance and time away from work, limited clinic hours, long waits, stockouts of consumables, and scarcity of trained providers reduce access. Weak referral and recall arrangements, fragmented registers, and lack of unique identifiers contribute to attrition after a positive screen [8,7]. Together, these barriers explain why programmes that rely on passive and opportunistic uptake rarely achieve population level coverage without proactive invitation and supportive system design.

### Interventions that work: evidence from low- and middle-income countries and Nigeria

#### Targeted health education and invitation or recall

Structured, culturally sensitive education delivered through community and workplace channels improves knowledge and intent to screen. Behavioural strategies such as commitment prompts, appointment reminders and default scheduling can further increase completion [7,2]. Where workplace communication systems exist, coordinated campaigns can be timed to align with screening availability and staff rosters.

#### HPV self-sampling as a demand side lever

Randomised and observational evidence in Nigeria and other low- and middle-income countries indicates that self-collected HPV samples are acceptable, feasible and increase screening uptake compared with clinician collected sampling [10,2]. Self-sampling reduces time and embarrassment costs and is operationally compatible with workplace distribution plus drop off or courier return. Programmes must ensure reliable laboratory logistics, results communication, and linkage to triage and treatment [2].

#### Screen and treat models to reduce attrition

Same day treatment after a positive screen, when clinically appropriate, reduces loss to follow up. Operational success depends on trained providers, functional equipment for thermal ablation or cryotherapy, clear eligibility criteria, and referral for ineligible lesions [2,6]. Embedding treatment sessions within planned outreach days or dedicated clinic blocks helps maintain throughput.

#### Service integration for reach and equity

Integrating screening into maternal and child health, family planning, and HIV clinics increases contact opportunities and can leverage existing community trust and recall infrastructure [2]. Workplace health services are an extension of this strategy, offering a high yield setting for reaching midlife women who may otherwise struggle to attend primary care during working hours.

#### Data systems, SMS recalls and quality improvement routines

Simple, standardised registers and unique client identifiers underpin reliable tracking of invitations, screening, positivity, treatment and recalls. SMS reminders before appointments and follow up prompts after positive results are low cost and scalable. Routine quality improvement huddles, weekly or monthly and run charts support problem solving on bottlenecks that include missed appointments, specimen transport delays and equipment downtime, and help to sustain gains [13,2].

### Why a quality improvement approach suits Nigeria workplace clinics

The gap between policy and delivery in cervical cancer control is chiefly an implementation problem, and quality improvement is designed for exactly this space [12,14]. PDSA cycles allow frontline teams to test context specific changes in small batches, for example a new email subject line, a script for nurse led invitations, default appointment slots, or simplified booking, and to adopt, adapt or abandon changes based on real time data. Quality improvement emphasises process measures such as the proportion of invited women who schedule an appointment within seven days, outcome measures such as completed Pap or HPV tests per 100 eligible women per quarter, and balancing measures such as nurse overtime hours and waiting time [13]. The discipline of visualising data over time through run charts and control charts helps distinguish common cause fluctuation from true improvement and aligns staff around shared goals.

Workplace clinics such as the Central Bank of Nigeria Clinic have clear communication channels, predictable clinic schedules and administrative support, all conducive to running sequential PDSA cycles efficiently. Moreover, lessons learned in these controlled environments can inform adaptation for community primary health centres that face higher variability in patient flow and fewer communication tools. Demonstrating that modest and low-cost changes, including timed lectures, targeted emails, SMS reminders, simplified booking and audit and feedback, can deliver meaningful gains in screening within three months offers practical value for health managers working with constrained budgets [6,5].

### Evidence gaps and contribution of the present project

Despite a growing literature on HPV vaccination and screening in Nigeria, several gaps remain. First, few published reports describe workplace-based quality improvement interventions with quantitative before and after changes in both knowledge and screening uptake. Second, while several Nigerian studies document low awareness and screening prevalence, fewer link demand creation directly to completed screening using routine clinic data and statistical testing of knowledge gains [8,7]. Third, although national and WHO standards endorse HPV DNA testing as the preferred modality, many facilities still operate with Pap or visual inspection with acetic acid because of resource constraints. Pragmatic improvement stories from such settings are important for near term mortality reduction [2,6]. Finally, there is limited synthesis on how to operationalise invitation, screening, treatment and recall within existing data systems to ensure traceability and timely feedback to teams [13,2].

The Central Bank of Nigeria Clinic initiative addresses these gaps by mobilising a defined and reachable population through a structured education event and multi-channel messaging, by measuring knowledge change with before and after surveys, by using routine clinic records to quantify screening uptake over comparable periods, and by embedding the intervention within a quality improvement frame that is intentionally low cost and scalable. The focus on a workplace clinic complements community strategies and offers a blueprint for other occupational health services in Nigeria.

### Conceptual framing and theory of change

Our theory of change posits that if evidence-based education tailored to local misconceptions is delivered through high reach channels, and logistical frictions are reduced through simple default appointments and reminders, then more eligible women will present for screening, and a higher proportion of screen positive women will receive timely evaluation and treatment. Enablers include trained providers, clear eligibility criteria for same day treatment, and minimal administrative steps at the point of care [2,6]. Quality improvement routines that include PDSA, run charts and feedback provide the mechanism for iterative refinement and sustainability [12,13]. In the medium term, the model can incorporate HPV self-sampling when feasible, which is likely to yield a further step change in coverage while preserving the same backbone for invitations, results and recalls [10,2].

## Methods

### Study design and setting

This quality improvement project was conducted in November 2024 at the Central Bank of Nigeria in Abuja, Nigeria.

The intervention was developed using quality improvement principles, focusing on demand creation through education and reduction of logistical barriers to screening.

### Participants

All CBN staff were invited to attend the educational hybrid session; there were accommodations for both virtual and physical attendance. Invites and reminders were sent through paper flyers, emails, social media (Viva engage), and electronic bulletin board. All attendees were included in knowledge assessment, but men were excluded from pap smear uptake data.

### Intervention

The intervention comprised of a structured educational session which covered cervical cancer epidemiology, risk factors, HPV infection, signs and symptoms, primary prevention, screening guidelines and techniques, treatment and common myths and cultural misconceptions. The session was deliberately created to be a safe, nonjudgmental space for participants to ask questions.

In addition, system-level changes were implemented at the clinic level. We helped create a simplified scheduling system for patients to make pap smear appointments through a clear pathway for the visit. In addition, we, engaged providers around screening guidelines and follow up recommendations.

### Data collection and outcomes

Participants completed pre- and post-intervention questionnaires assessing knowledge across ten key items, along with demographic characteristics including age bracket, gender, educational level, and marital status. A QR code was displayed and printed for the session. Paper copies of the surveys were also printed for participants that had any technical issues.

Primary outcomes included change in knowledge and pap smear uptake numbers. Screening uptakes were gathered using routine clinic records for three months following the intervention.

### Ethics

IRB approval was obtained from the Central Bank of Nigeria. External institutional review board was not required as the project was classified as a quality improvement project and not human subjects’ research.

### Statistical analysis

Descriptive statistics summarized participant demographic characteristics. Pre- and post-intervention survey responses were compared using*** statistical tests, with statistical significance set as p<0.05. Pap smear uptake was compared descriptively between pre- and post-intervention periods.

## Results

### Participation and survey responses

A total of 188 staff attended the educational sessions. One hundred and twenty-four survey responses were analyzed, including 70 pre-intervention and 54 post-intervention responses. Figure 1 and Figure 2.

**Figure 1:**
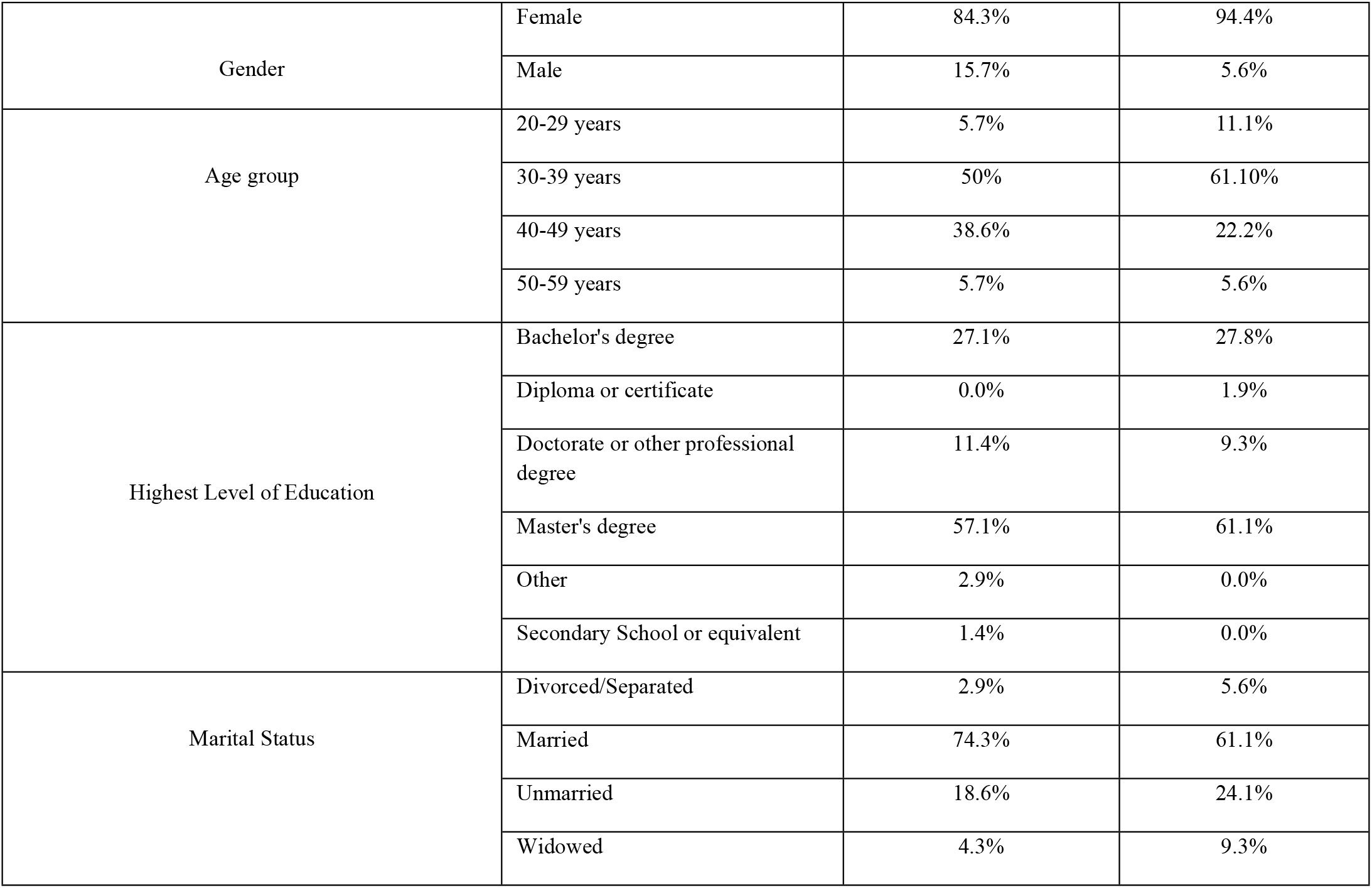
Demographics

**Figure 2:**
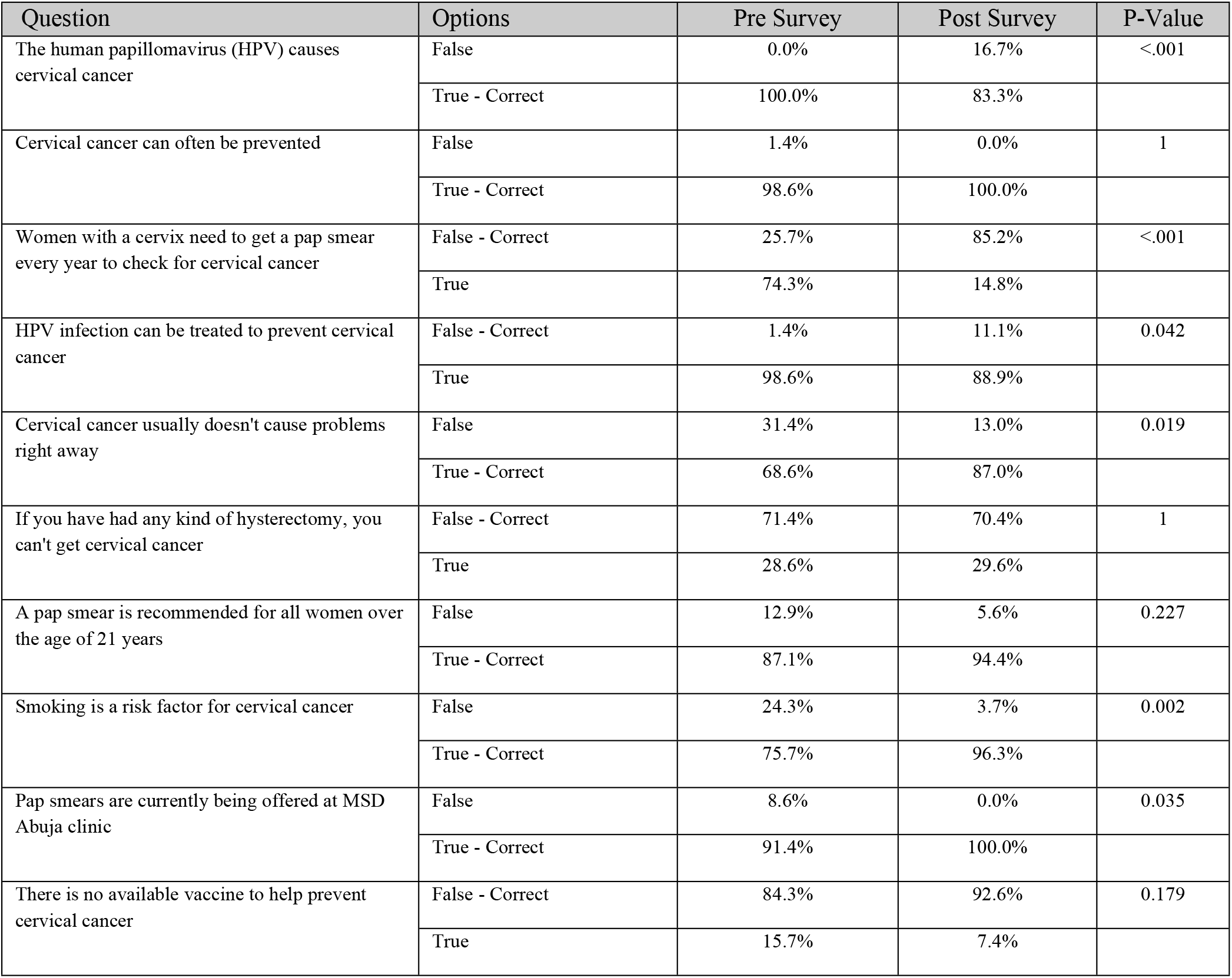
Results

### Knowledge outcomes

Knowledge improvement was observed in eight of the ten questionnaire items. Five knowledge domains demonstrated statistically significant improvement following the intervention (p<0·05) (Table 1).

**Table 1.**
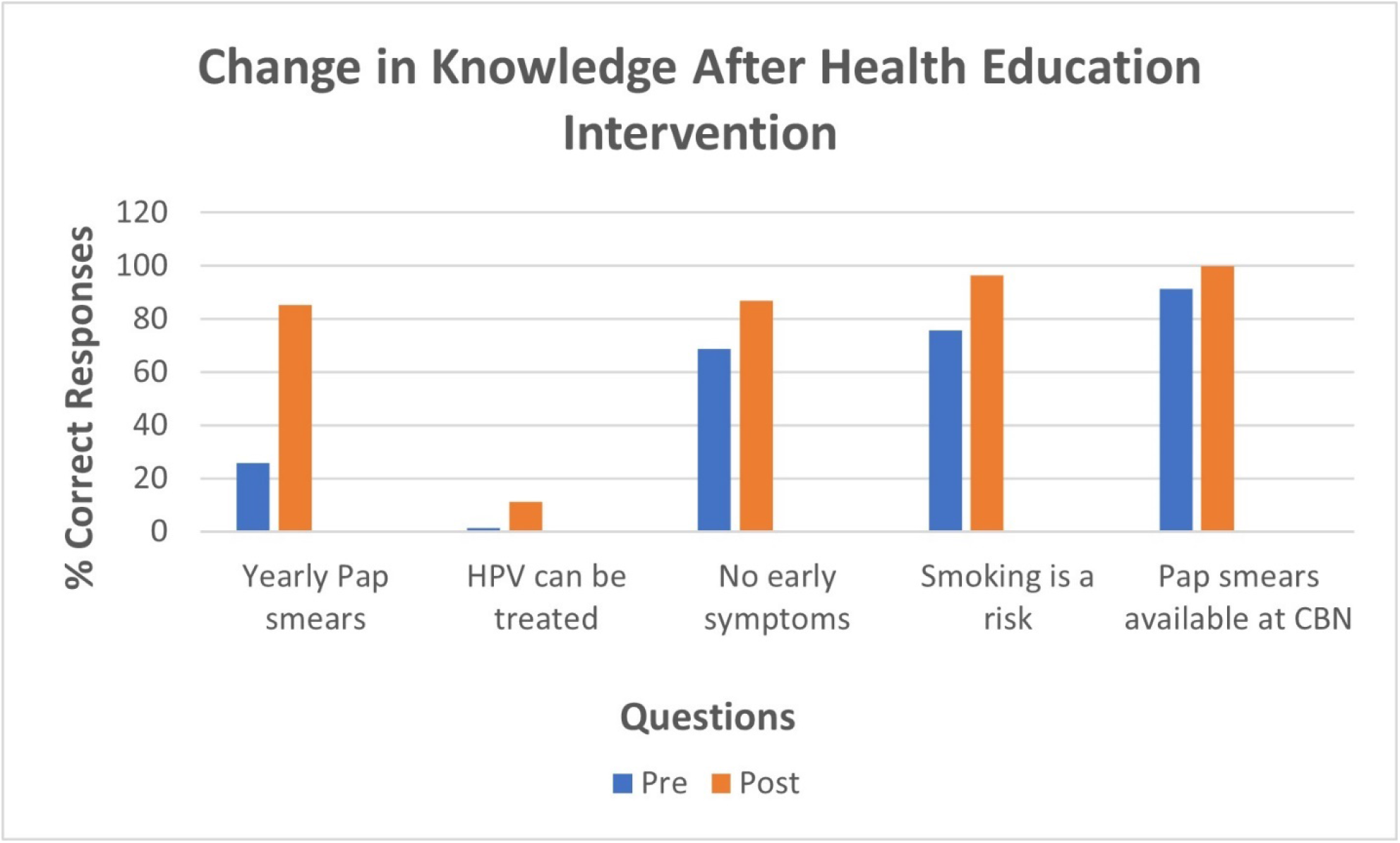

### Screening uptake

Pap smear uptake increased from five women screened in the three months prior to the intervention to 32 women screened in the three months following the intervention. Table 2

**Table 2.**

| Metric | Pre-Intervention | Post-Intervention |
| --- | --- | --- |
| Survey Respondents | 70 | 54 |
| Pap Smear Uptake | 5 | 32 |
| Knowledge Areas with significant gains | - | 5 |
| Total Attendees 188 |  |  |

### Participant feedback

Participants consistently reported that the educational sessions provided a safe and supportive environment to clarify misconceptions, discuss cultural beliefs, and ask questions about cervical cancer, HPV vaccination, and screening.

## Discussion

This quality improvement initiative demonstrated that target educational sessions focusing on common myths and cultural misconceptions fused with system-level changes are associated with significant improvements in cervical cancer knowledge and pap smear screening uptake within a short period. Including male counterparts in the educational session and knowledge assessment recognized the role they play in shaping health norms and decision-making within families and workplaces.

Educational interventions addressing myths, cultural beliefs, and knowledge gaps are increasingly recognized as critical to improving cervical cancer screening uptake in Nigeria and across Africa [6–9]. This intervention’s value extends beyond delivery of information. Creating a safe, nonjudgmental environment was essential in addressing and reducing stigma and fear, in enabling participants to discuss openly, and in translating the knowledge into action. Barriers to HPV vaccination and pap smear uptake were further addressed through creating a simplified scheduling system for the clinic which helped ease the process of getting screened and receiving the vaccine. There is emerging evidence supporting HPV self-swabs as a future strategy in similar settings [10,11].

### Implications for policy and practice

For policymakers and programme managers, three implications are salient. First, workplace clinics are an underused delivery channel for secondary prevention that can generate quick wins toward national targets at low marginal cost. Second, the combination of targeted education, proactive invitation and recall, and simplified pathways for screening and treatment is an immediately actionable bundle that aligns with WHO standards while accommodating local constraints [2,5]. Third, embedding light touch data systems and quality improvement routines enables continuous learning and scale up. The same dashboards that serve a single clinic can be federated across multiple facilities to support regional improvement collaboratives [13]. As Nigeria continues to expand HPV vaccination, parallel investments in adult screening and treatment implemented through quality improvement are essential to bend mortality curves within the current decade [4,1].

The elimination of cervical cancer as a public health problem will not be achieved by guidelines alone but by reliable and context appropriate delivery systems. Nigeria persistent gaps in awareness, screening coverage and timely treatment underscore the need for practical implementation strategies. Workplace clinics offer a tractable setting in which quality improvement methods can be applied to raise demand and throughput quickly. The present project contributes evidence that targeted education and multi-channel communication can improve knowledge and increase completed screening in the short term, providing a template for broader scale up while HPV testing capacity and referral networks mature.

### Limitations

The study’s short follow-up period limits conclusions regarding long-term sustainability. Screening uptake was assessed using aggregate clinic data rather than individual-level linkage to survey responses. Nevertheless, the magnitude and timing of the observed increase support a meaningful intervention effect.

## Conclusion

Educational interventions that are myth-focused, culturally sensitive, and rooted within an accommodating health system processes can significantly improve cervical cancer knowledge and screening uptake. Workplace health care centers or clinics are underutilized but provide a high-yield platform for secondary prevention. Applying quality improvement processes in such settings in conjunction with continued expansion of HPV vaccination and more advanced screening can offer an accessible route to aid in cervical cancer elimination in Nigeria.

## Data Availability

The data underlying this study are fully de-identified and do not contain any personal identifiers such as names or employee numbers. The dataset includes demographic and study-related variables. Data are available from the corresponding author upon reasonable request, and may be shared in a suitable public repository following publication, in accordance with institutional and ethical guidelines.

## Notes

### Competing Interest Statement

The authors have declared no competing interest.

### Funding Statement

The authors received no external funding for this work. The study was supported through institutional resources, including reimbursement of travel expenses by the residency program. The funders had no role in study design, data collection and analysis, decision to publish, or preparation of the manuscript.

### Author Declarations

This study was reviewed by the Institutional Review Board (IRB) of WellSpan Health and was determined to be exempt from full review. The project involved minimal risk and utilized de-identified data with no personal identifiers such as names or employee numbers. The study was conducted in accordance with institutional ethical guidelines.

